# Vascular Risk Factors, Biological Aging, and Cognitive Performance in a National Sample of Older U.S. Adults

**DOI:** 10.64898/2026.03.12.26348280

**Authors:** Jenny Jyoung Lee, Anisha Das, Tristin Yun, Annie J. Lee

## Abstract

Cardiovascular and cerebrovascular risk factors (CVRFs)—including hypertension, diabetes, heart disease, and stroke—are prevalent chronic conditions in older adults and major determinants of late-life cognitive decline. These conditions involve chronic inflammatory and metabolic processes that may accelerate biological aging, reflecting multisystem physiological decline beyond chronological age. We examined associations among CVRFs, accelerated biological aging, and cognitive performance and assessed whether biological aging mediates the association between CVRFs and cognitive performance overall and across race/ethnicity and sex. We analyzed data from 2,384 U.S. adults aged 60 years and older in the National Health and Nutrition Examination Survey 2011–2014. CVRFs were defined using clinical measurements and self-reported diagnoses. Biological aging was quantified using the PhenoAge algorithm derived from blood-based clinical biomarkers. Cognitive performance was assessed using composite scores of memory, executive function, and processing speed. Weighted linear regression and causal mediation analyses were conducted overall and stratified by race/ethnicity and sex. All CVRFs were associated with accelerated biological aging, with diabetes demonstrating the strongest association (0.76 SD higher PhenoAge acceleration; 95% CI: 0.67–0.85). CVRFs were associated with lower cognitive performance, with stroke showing the largest association (β = −0.317; 95% CI: −0.471 to −0.165). Accelerated biological aging mediated these associations, accounting for 88.5% of the diabetes association and 13.7%–27.2% for other CVRFs. Associations and mediation effects varied across racial/ethnic and sex groups, with mediation more consistent among Non-Hispanic Whites and females. Accelerated biological aging represents an important link between cardiometabolic risk to cognitive performance in older adults.

## Introduction

Cardiovascular and cerebrovascular risk factors (CVRFs)—including hypertension, diabetes, heart disease, and stroke—are among the most prevalent chronic conditions in older adults and are major determinants of late-life cognitive decline^1–4^. Although these conditions are modifiable through prevention and clinical management^5^, their broader contribution to systemic biological aging remains incompletely understood. CVRFs are characterized by chronic inflammation, metabolic dysregulation, and endothelial dysfunction^6^ —processes that are fundamental to the biology of aging. These shared mechanisms suggest that CVRFs may contribute to cognitive impairment, in part, through acceleration of systemic biological aging.

Biological aging reflects progressive multisystem physiological dysregulation that accompanies advancing age and contributes to heterogeneity in health outcomes among older adults^7^. Variation in the pace of biological aging is detectable by midlife and predict morbidity and mortality independent of chronological age^8,9^. Experimental studies in model organisms demonstrate that core aging pathways are modifiable, as interventions targeting fundamental mechanisms of aging extend healthy lifespan^7^. In human populations, accelerated biological aging has been associated with poorer cognitive performance and greater cognitive decline^8^.

Recent advances in algorithm-based measures have enabled quantification of biological aging using molecular and clinical biomarker data^10,11^. Epigenetic clocks based on DNA methylation patterns show robust associations with morbidity and mortality^12,13^, while clinical laboratory–based measures provide scalable, population-relevant alternatives^11^. Among these, PhenoAge integrates chronological age with routinely collected clinical biomarkers to estimate phenotypic age and mortality risk^11^. PhenoAge predicts morbidity and mortality and is increased among individuals with cardiometabolic conditions, including hypertension, diabetes, heart disease, and stroke^14^. However, vascular risk, biological aging, and cognition have largely been examined in isolation. Whether accelerated biological aging mediates the association between CVRF burden and cognitive performance remains unclear.

Clarifying the role of biological aging in linking cardiometabolic risk to cognitive aging has important public health implications, given persistent disparities across racial/ethnic and sex groups. Hypertension and diabetes are more common among Non-Hispanic Black and Hispanic adults than among Non-Hispanic White adults^15,16^, and dementia risk is approximately 1.5 to 2 times higher in these populations^17^. Determining whether accelerated biological aging contributes to these differences may clarify mechanisms underlying disparities in late-life cognitive impairment and inform strategies to reduce cognitive decline.

In this study, we examined the associations among major CVRFs (hypertension, diabetes, heart disease, and stroke), accelerated biological aging, and cognitive performance in a nationally representative sample of older U.S. adults from the National Health and Nutrition Examination Survey (NHANES). We further tested whether accelerated biological aging mediates the associations between CVRF burden and cognitive performance using causal mediation analysis and evaluated heterogeneity across racial/ethnic and sex groups.

## Methods

### Study Design and Population

The National Health and Nutrition Examination Surveys (NHANES) study is a long-standing federal survey that integrates interview, examination, and laboratory components to provide nationally representative information on the health of U.S. adults and children^18^. NHANES has been conducted in independent 2-year cross-sectional cycles since 1999, using a multistage probability sampling strategy with oversampling of specific demographic subgroups to improve the accuracy of subgroup estimates. Data collection begins with an in-home interview in which trained staff obtain demographic, socioeconomic, medical, and behavioral information. Participants are subsequently examined in a mobile examination center, where certified technicians administer standardized physical measurements, collect biological specimens, and conduct dietary and laboratory assessments following protocols established by the National Center for Health Statistics. Extensive documentation on survey operations, instrumentation, and quality-control procedures is publicly available through NHANES^18^. All study protocols were approved by the National Center for Health Statistics Research Ethics Review Board, and written informed consent was obtained from all participants.

We focused on the 2011–2014 NHANES^19^ cycles because these were the only survey years in which the cognitive assessments and biomarker data required for this study were both available and comparable. Among the 11,329 individuals examined during this period, we included participants who met the following criteria: (1) aged ≥60 years, as cognitive testing was restricted to this age group; (2) availability of sociodemographic information (sex, race/ethnicity, educational attainment, marital status, and income-to-poverty ratio); (3) cognitive data from three standardized neuropsychological assessments (CERAD Delayed

Recall Test, the Animal Fluency Test, and the Digit Symbol Substitution Test); (4) laboratory measurements for eight clinical biomarkers required to estimate biological age; and (5) information on four cardio-cerebrovascular risk factors (hypertension, diabetes, heart disease, and stroke). After applying these criteria, the final analytic sample included 2,384 participants (**Figure 1**).

**Figure 1.**
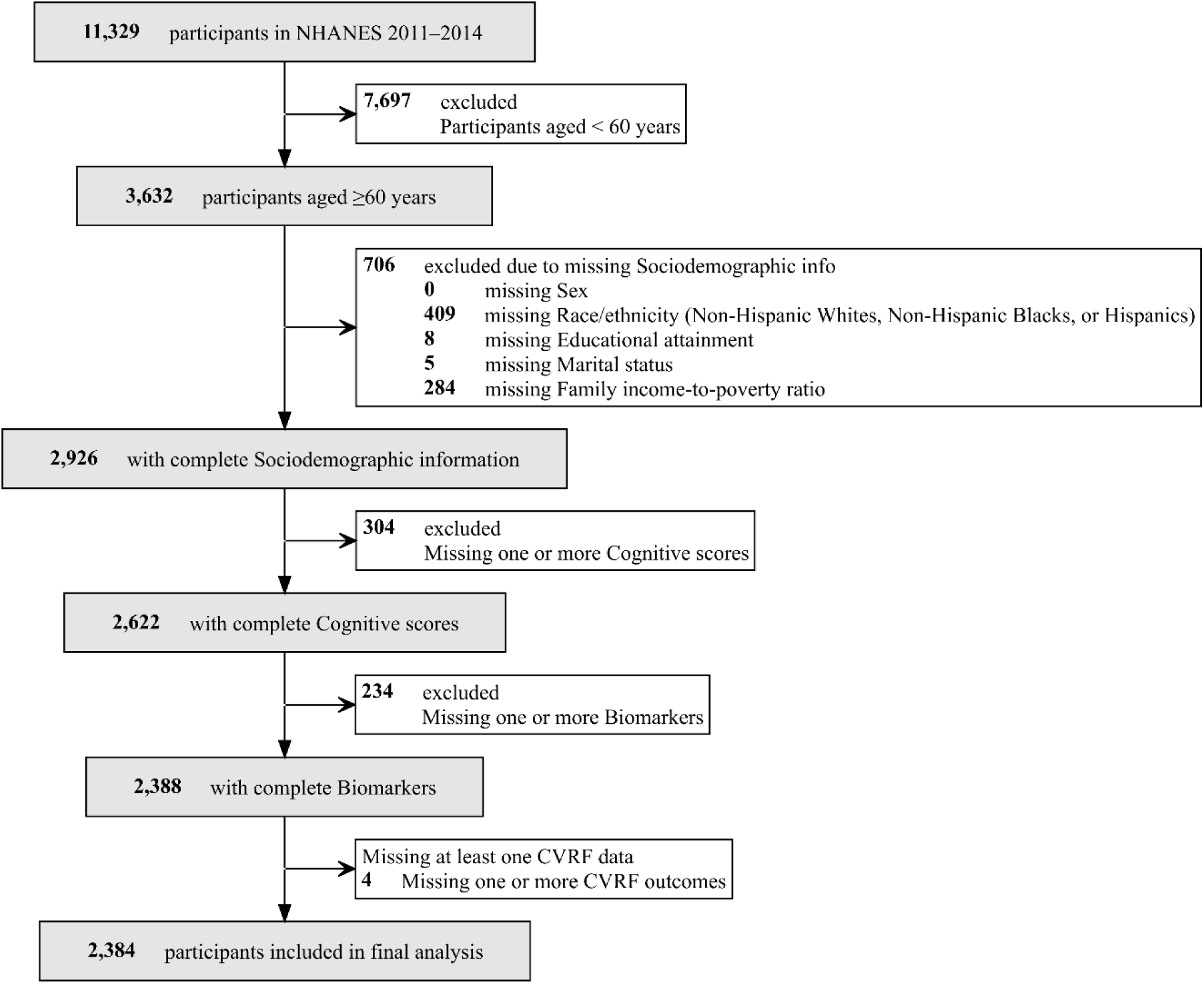
Flow diagram of participant selection.

### Cardio-cerebrovascular Risk Factors

Cardio-cerebrovascular risk factors—hypertension, diabetes, heart disease, and stroke—were defined using harmonized information from self-reported medical history and clinical measurements^20^. Hypertension was classified based on a self-reported history of hypertension, systolic blood pressure ≥140 mmHg, or diastolic blood pressure ≥90 mmHg. Diabetes was identified by self-reported diagnosis or glycated hemoglobin A1c (HbA1c) ≥7%. Heart disease was defined as a self-reported history of congestive heart failure, coronary heart disease, angina, or myocardial infarction. Stroke status was determined from a self-reported history of physician-diagnosed stroke.

### Biological Aging

Biological age was estimated using the PhenoAge algorithm^11^, a validated biomarker-based measure of aging derived from clinical blood chemistry data and chronological age to estimate an individual’s mortality risk. PhenoAge was selected as a well-validated biological aging measure compatible with NHANES and originally developed using NHANES III (1988–1994) laboratory data. PhenoAge is strongly predictive of chronic disease, functional decline, and mortality^11,14,21^, and outperforms alternative blood chemistry–based biological age algorithms and several DNA methylation–based aging measures^14,22^. The PhenoAge model was developed using elastic-net regression to predict mortality risk from chronological age and a panel of clinical biomarkers. Model coefficients were trained in NHANES III participants aged 20–85 years to establish biomarker–age relationships across the adult life course and were subsequently applied to the present study population. PhenoAge was computed using eight available biomarkers: albumin, alkaline phosphatase, creatinine, glycated hemoglobin (HbA1c), white blood cell count, lymphocyte percentage, mean cell volume, and red cell distribution width. PhenoAge was calculated using the published algorithm as implemented in the BioAge R package^23^.

To quantify biological aging relative to chronological age, we calculated PhenoAge acceleration as the residual from a linear regression of PhenoAge on chronological age. The residuals were standardized to a mean of 0 and a standard deviation of 1, and the resulting standardized PhenoAge acceleration served as the primary measure of accelerated biological aging in subsequent analyses. Positive values indicate PhenoAge higher than expected for a given chronological age (i.e., accelerated biological aging), whereas negative values indicate slower or delayed biological aging.

### Cognitive Performance

Cognitive performance was assessed using three standardized neuropsychological tests administered to NHANES participants aged ≥60 years during 2011–2014^18^: the Consortium to Establish a Registry for Alzheimer’s Disease (CERAD) Delayed Recall Test, the Animal Fluency Test (AFT), and the Digit Symbol Substitution Test (DSST). The CERAD Delayed Recall Test evaluates episodic memory by asking participants to recall a list of 10 previously presented words after a brief delay (score range: 0–10). The AFT measures verbal fluency and executive functioning by instructing participants to name as many animals as possible within 60 seconds, with one point awarded per correct response. The DSST, a subtest of the Wechsler Adult Intelligence Scale–III, assesses processing speed, sustained attention, and working memory by requiring participants to match symbols to numbers using a reference key within 120 seconds. Raw scores from each test were standardized to z-scores and a global cognitive score was then derived by averaging the three standardized test scores, providing a composite measure of overall cognitive function.

### Demographic and Socioeconomic Covariates

Sociodemographic information was obtained from the NHANES household interview and included age, sex, and race/ethnicity, categorized as Non-Hispanic White, Non-Hispanic Black, and Hispanic^18,24^. Socioeconomic variables included educational attainment (less than high school; high school or some college; bachelor’s degree or higher), marital status (married or cohabitating; divorced, widowed, or separated; never married), and the income-to-poverty ratio. Consistent with thresholds used in the Affordable Care Act, the income-to-poverty ratio was classified as low (≤1.00), middle (1.01–3.99), and high (≥4.00)^25,26^.

### Statistical Analyses

All analyses accounted for the complex sampling design of NHANES, including survey strata, primary sampling units, and sampling weights, to generate nationally representative estimates. In accordance with NHANES analytic guidelines for biomarker data collected in the Mobile Examination Center (MEC), MEC examination weights were applied in all weighted analyses. When combining data from the 2011–2012 and 2013–2014 cycles, the 2-year MEC weights were divided by two to derive appropriate weights for the combined 4-year analytic period.

Participant characteristics were summarized using means and standard deviations (SD) for continuous variables and percentages for categorical variables. Statistical differences by race/ethnicity and sex were assessed using analysis of variance (ANOVA) for continuous variables and Chi-Square for categorical variables. Statistical significance was defined as a two-sided p < 0.05.

To evaluate the role of biological aging in the association between CVRFs and cognitive performance, we estimated a series of weighted linear regression models examining three primary relationships: (1) associations between each CVRF and biological aging, (2) associations between biological aging and cognitive performance, and (3) associations between each CVRF and cognitive performance. To assess whether biological aging mediated the effect of CVRFs on cognitive function, we employed causal mediation analysis framework^27,28^. These analyses decomposed the total effect of each CVRF on cognitive performance into a direct effect (not operating through biological aging) and an indirect effect mediated through biological aging (**Figure 3**). We report the Average Causal Mediation Effect (ACME), Average Direct Effect (ADE), total effect, and proportion mediated, along with 95% confidence intervals estimated using a quasi-Bayesian approximation with 1,000 simulations. Mediation analyses were conducted using mediation R package^29^. Covariates were included in two sequential models. Model 1 adjusted for demographic factors (sex, race/ethnicity, and NHANES survey cycle). Model 2 additionally adjusted for socioeconomic factors (educational attainment, income-to-poverty ratio, and marital status). All analyses were conducted for the full sample and separately stratified by race/ethnicity and sex to assess subgroup differences. All statistical analyses were performed using R (version 4.4.2).

## Results

### Study Population Characteristics

The analytic sample included 2,384 older adults with a mean chronological age 73.79 (± 5.61) years and a mean global cognitive z-score of 0.01 (± 0.81) (**Table 1**). The sample was 51.5% female, and participants self-identified as Non-Hispanic White (54.6%), Non-Hispanic Black (24.7%), or Hispanic (20.7%). CVRFs were common, including hypertension (79.6%), diabetes (23.7%), heart disease (18.1%), and stroke (7.7%). Socioeconomic characteristics, including educational attainment, marital status, and family income-to-poverty ratio, differed significantly across race/ethnicity and sex. The mean PhenoAge was 72.77 (± 8.38) years, approximately one year lower than the mean chronological age (73.79 ± 5.61 years). Chronological age and PhenoAge were moderately correlated (r=0.692; **Figure S1**), with similar correlations across sex and race/ethnicity strata (r=0.666–0.703).

**Table 1.** Characteristics of the Study Participants, NHANES 2011–2014 (n = 2,384).

|  | Overall | Non-Hispanic White | Non-Hispanic Black | Hispanic | p | Male | Female | p |
| --- | --- | --- | --- | --- | --- | --- | --- | --- |
| **n** | 2,384 | 1,302 | 588 | 494 |  | 1,157 | 1,227 |  |
| **Race/Ethnicity** |  |  |  |  |  |  |  |  |
| Non-Hispanic White | 1,302 (54.6) | 1,302 (100.0) | 0 (0.0) | 0 (0.0) | - | 613 (53.0) | 689 (56.2) | 0.221 |
| Non-Hispanic Black | 588 (24.7) | 0 (0.0) | 588 (100.0) | 0 (0.0) |  | 302 (26.1) | 286 (23.3) |  |
| Hispanic | 494 (20.7) | 0 (0.0) | 0 (0.0) | 494 (100.0) |  | 242 (20.9) | 252 (20.5) |  |
| **Sex** |  |  |  |  |  |  |  |  |
| Male, n(%) | 1,157 (48.5) | 613 (47.1) | 302 (51.4) | 242 (49.0) | 0.221 | 1157 (100.0) | 0 (0.0) | - |
| Female, n(%) | 1,227 (51.5) | 689 (52.9) | 286 (48.6) | 252 (51.0) |  | 0 (0.0) | 1,227 (100.0) |  |
| **Educational Attainment** |  |  |  |  |  |  |  |  |
| Under high school, n (%) | 290 (12.2) | 60 (4.6) | 60 (10.2) | 170 (34.4) | <2.2e-16 | 160 (13.8) | 130 (10.6) | 0.049 |
| High school/Some college, n (%) | 910 (38.2) | 450 (34.6) | 293 (49.8) | 167 (33.8) |  | 428 (37.0) | 482 (39.3) |  |
| Bachelors' degree/Higher, n (%) | 1,184 (49.7) | 792 (60.8) | 235 (40.0) | 157 (31.8) |  | 569 (49.2) | 615 (50.1) |  |
| **Marital Status** |  |  |  |  |  |  |  |  |
| Married/Cohabiting, n (%) | 1,342 (56.3) | 775 (59.5) | 269 (45.7) | 298 (60.3) | 1.3e-09 | 799 (69.1) | 543 (44.3) | <2.2e-16 |
| Divorced/Widowed/Separated, n (%) | 911 (38.2) | 474 (36.4) | 263 (44.7) | 174 (35.2) |  | 294 (25.4) | 617 (50.3) |  |
| Never married, n (%) | 131 (5.5) | 53 (4.1) | 56 (9.5) | 22 (4.5) |  | 64 (5.5) | 67 (5.5) |  |
| **Family income-to-poverty Ratio** |  |  |  |  |  |  |  |  |
| Below federal poverty level, n (%) | 407 (17.1) | 142 (10.9) | 126 (21.4) | 139 (28.1) | <2.2e-16 | 163 (14.1) | 244 (19.9) | 1.35e-04 |
| Middle Income, n (%) | 1,358 (57.0) | 739 (56.8) | 334 (56.8) | 285 (57.7) |  | 663 (57.3) | 695 (56.6) |  |
| High Income, n (%) | 619 (26.0) | 421 (32.3) | 128 (21.8) | 70 (14.2) |  | 331 (28.6) | 288 (23.5) |  |
| **CVRFs** |  |  |  |  |  |  |  |  |
| Hypertension, n (%) | 1,898 (79.6) | 1,029 (79.0) | 496 (84.4) | 373 (75.5) | 0.001 | 906 (78.3) | 992 (80.8) | 0.137 |
| Diabetes, n (%) | 566 (23.7) | 251 (19.3) | 173 (29.4) | 142 (28.7) | 1.35e-07 | 292 (25.2) | 274 (22.3) | 0.105 |
| Heart Disease, n (%) | 432 (18.1) | 280 (21.5) | 85 (14.5) | 67 (13.6) | 1.44e-05 | 247 (21.3) | 185 (15.1) | 8.87e-05 |
| Stroke, n (%) | 184 (7.7) | 111 (8.5) | 55 (9.4) | 18 (3.6) | 0.001 | 81 (7.0) | 103 (8.4) | 0.231 |
| **Cognitive Score, mean (SD)** | 0.01 (0.81) | 0.20 (0.80) | -0.24 (0.76) | -0.20 (0.77) | <2.2e-16 | -0.07 (0.78) | 0.09 (0.83) | 1.33e-06 |
| **NHANES Wave, n (%)** |  |  |  |  |  |  |  |  |
| 2011 - 2012 | 1,067 (44.8) | 541 (41.6) | 307 (52.2) | 219 (44.3) | 8.87e-05 | 529 (45.7) | 538 (43.8) | 0.379 |
| 2013 - 2014 | 1,317 (55.2) | 761 (58.4) | 281 (47.8) | 275 (55.7) |  | 628 (54.3) | 689 (56.2) |  |
| **Age and PhenoAge, mean (SD)** |  |  |  |  |  |  |  |  |
| Age | 73.79 (5.61) | 74.60 (5.44) | 73.15 (5.60) | 72.42 (5.72) | 7.13e-15 | 73.81 (5.65) | 73.77 (5.59) | 0.857 |
| PhenoAge | 72.77 (8.38) | 73.69 (8.30) | 72.32 (8.33) | 70.90 (8.32) | 6.64e-10 | 74.19 (8.50) | 71.44 (8.04) | 6.62e-16 |
| PhenoAge Acceleration | 0.00 (6.05) | 0.08 (5.92) | 0.21 (6.21) | -0.46 (6.21) | 0.157 | 1.40 (6.04) | -1.32 (5.76) | <2.2e-16 |
| Standardized PhenoAge Acceleration | 0.00 (1.00) | 0.01 (0.98) | 0.03 (1.03) | -0.08 (1.03) | 0.157 | 0.23 (1.00) | -0.22 (0.95) | <2.2e-16 |

When stratified by race/ethnicity, mean chronological age and PhenoAge differed significantly across groups (p < 0.001), with Non-Hispanic White participants exhibiting highest mean chronological and biological ages. CVRF prevalence differed across groups, with the highest prevalence of hypertension (84.4%), diabetes (29.4%), and stroke (9.4%) observed among Non-Hispanic Black participants, while heart disease was most prevalent among Non-Hispanic White participants (21.5%) and least prevalent among Hispanic participants (13.6%) (**Table 1**). Mean cognitive scores differed significantly across racial/ethnic groups (p < 0.001), with Non-Hispanic White participants showing higher scores than Non-Hispanic Black and Hispanic participants (0.20 vs. −0.24 vs. −0.20, p < 0.001). In contrast, PhenoAge acceleration did not differ significantly across racial/ethnic groups (p = 0.157).

When stratified by sex, chronological age did not differ between males and females (p = 0.857); however, males had higher mean PhenoAge than females (74.19 vs. 71.44, p < 0.001). Mean cognitive scores also differed by sex (p < 0.001), with females demonstrating higher scores than males (0.09 vs. −0.07). PhenoAge acceleration was significantly higher in males than females (1.40 ± 6.04 vs. −1.32 ± 5.76, p < 0.001). Hypertension, diabetes, and stroke prevalence did not differ by sex, whereas heart disease was more prevalent among males than females (21.3% vs. 15.1%, p < 0.001) (**Table 1**).

### Associations between CVRFs and biological aging

Cardio-cerebrovascular risk factors were consistently associated with accelerated biological aging in demographic-adjusted models, and these associations were modestly attenuated but remained statistically significant after additional adjustment for socioeconomic factors (**Figure 2** and **Table S1**). In fully adjusted models, participants with diabetes exhibited the strongest association with accelerated biological aging, corresponding to a 0.76 standard deviation (SD) higher standardized PhenoAge acceleration compared with those without diabetes (95% CI: 0.67–0.85). Heart disease was associated with a 0.50 SD increase (95% CI: 0.40–0.59), followed by stroke (0.32 SD; 95% CI: 0.17–0.46) and hypertension (0.34 SD; 95% CI: 0.26–0.43). Across race/ethnicity- and sex-stratified analyses, the direction and magnitude of associations were broadly consistent. However, the association between stroke and accelerated biological aging did not reach statistical significance among Non-Hispanic Black and Hispanic participants (**Table S1**).

**Figure 2.**
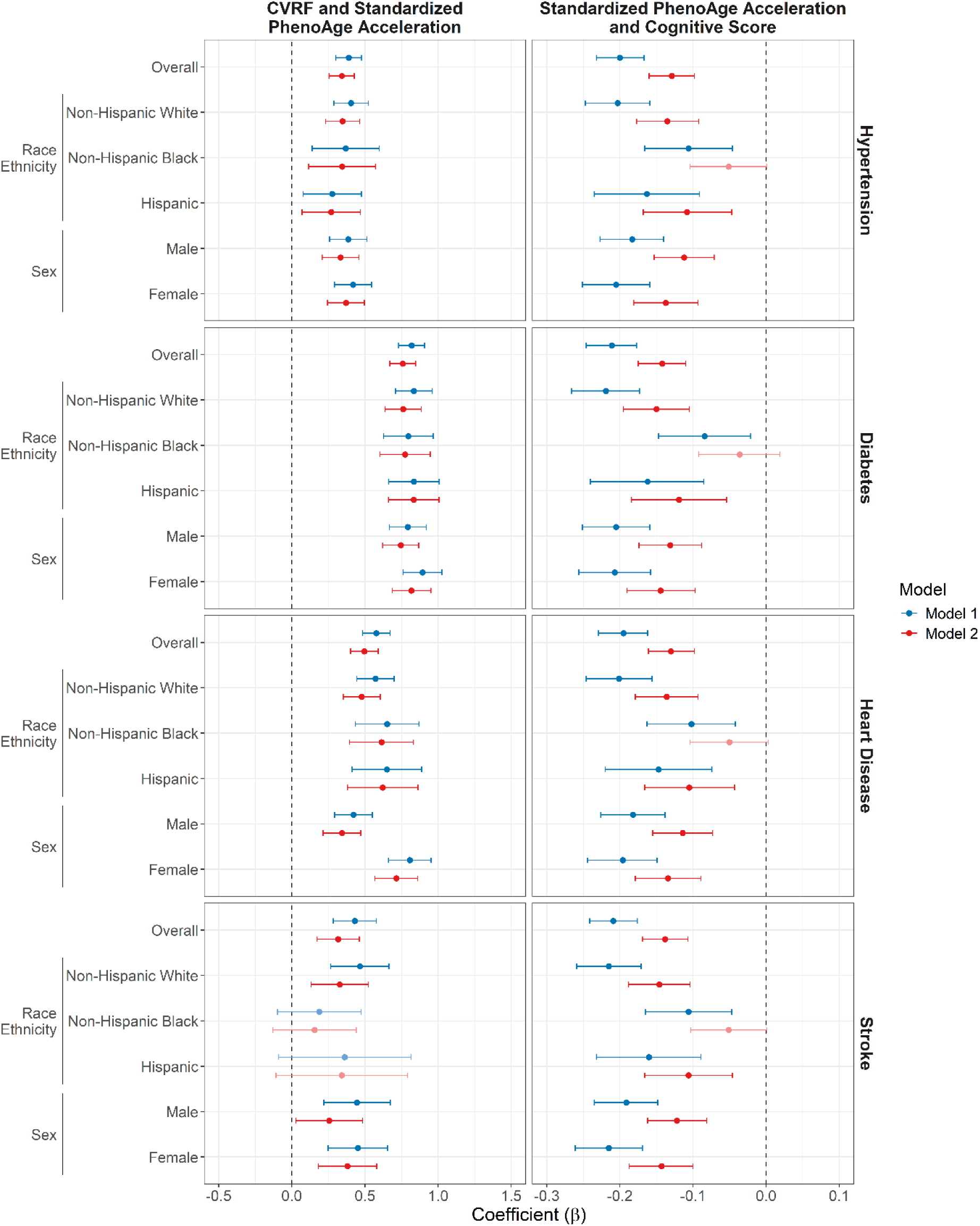
Associations of CVRFs with standardized PhenoAge acceleration and cognitive performance, overall and by race/ethnicity and sex, NHANES 2011–2014. Regression coefficients (β) and 95% confidence intervals from weighted linear regression models. Left panels show associations between CVRFs and standardized PhenoAge acceleration; right panels show associations between standardized PhenoAge acceleration and cognitive performance. Model 1 adjusts for demographic factors. Model 2 adjusts for demographic and socioeconomic factors.

**Figure 3.**
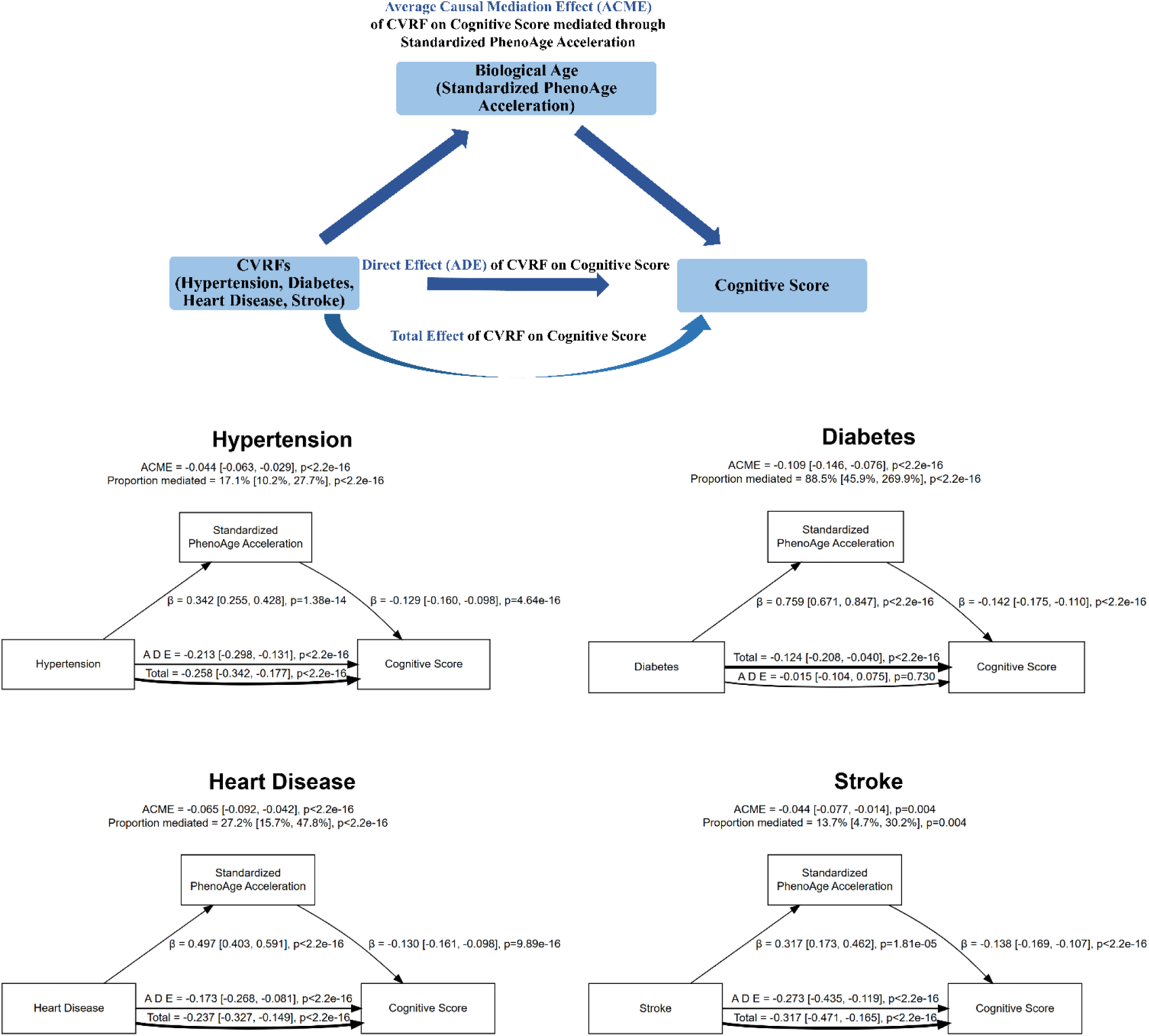
Mediation models linking CVRFs to cognitive performance through standardized PhenoAge acceleration, NHANES 2011–2014. Regression coefficients are shown for associations between CVRFs and standardized PhenoAge acceleration and between standardized PhenoAge acceleration and cognitive performance. Estimates of the average causal mediation effect (ACME), average direct effect (ADE), and total effect are presented. Models adjust for demographic and socioeconomic factors.

### Associations between CVRFs and cognitive performance

CVRFs were significantly associated with lower cognitive performance in the overall sample (total effects; **Figure 4** and **Table S2**). In fully adjusted models, stroke was associated with the largest reduction in cognitive scores (β = −0.317; 95% CI: −0.471 to −0.165), followed by hypertension (β = −0.258; 95% CI: −0.342 to −0.177), heart disease (β = −0.237; 95% CI: −0.327 to −0.149), and diabetes (β = −0.124; 95% CI: −0.208 to −0.040).

**Figure 4.**
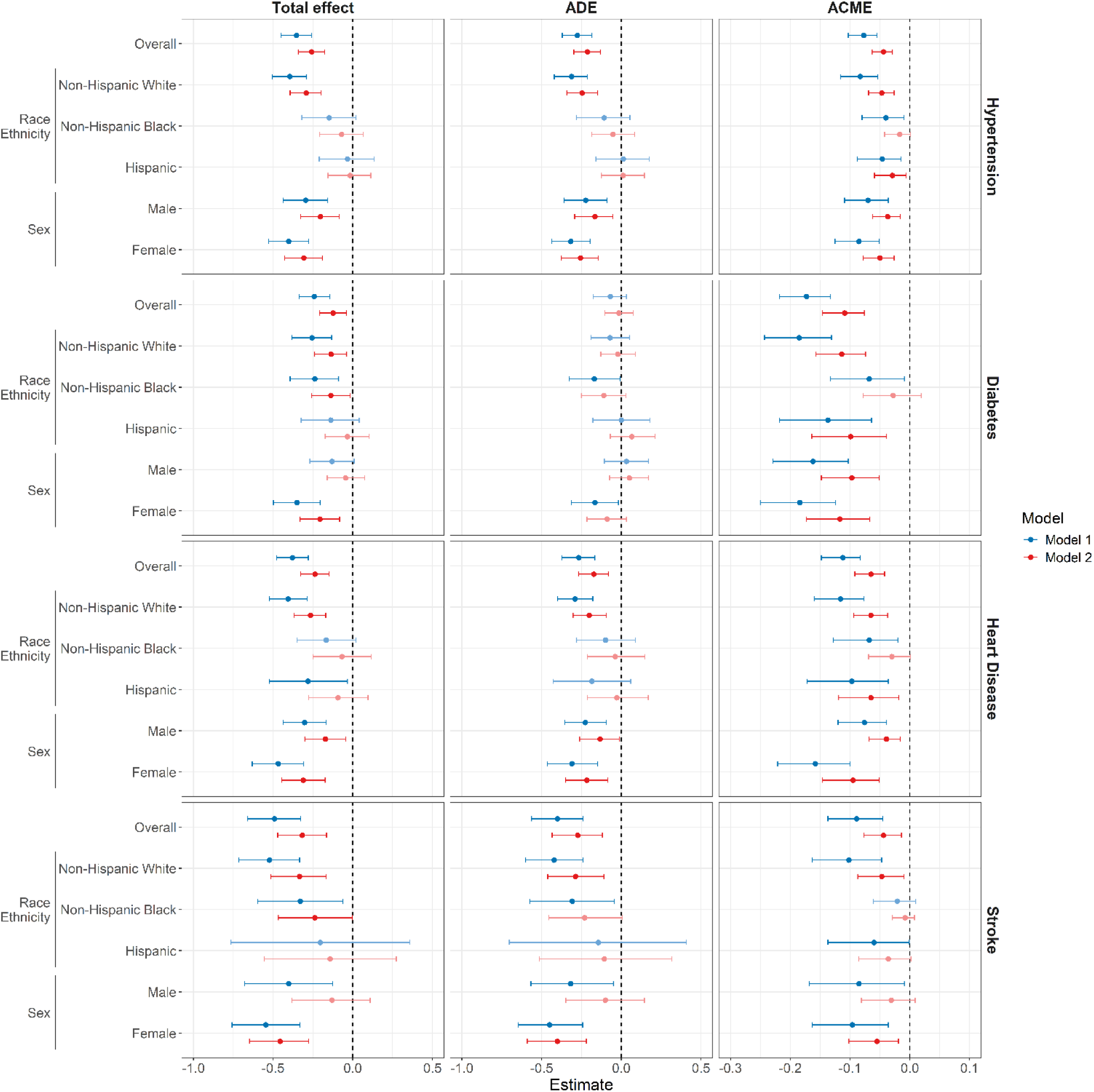
Mediation analysis of associations between CVRFs and cognitive performance via standardized PhenoAge acceleration, overall and stratified by race/ethnicity and sex, NHANES 2011–2014. Panels display total effects, average direct effects (ADE), and average causal mediation effects (ACME). Points represent regression estimates and bars indicate 95% confidence intervals. Model 1 adjusts for demographic factors. Model 2 additionally adjusts for socioeconomic factors.

Associations differed substantially by race/ethnicity (**Figure 4** and **Table S2**). Among Non-Hispanic White participants, all four CVRFs were associated with lower cognitive performance in fully adjusted models, including stroke (β = −0.334; 95% CI: −0.514 to −0.168), hypertension (β = −0.292; 95% CI: −0.393 to −0.198), heart disease (β = −0.265; 95% CI: −0.369 to −0.169), and diabetes (β = −0.136; 95% CI: −0.239 to −0.038). Among Non-Hispanic Black participants, significant associations persisted for stroke (β = −0.238; 95% CI: −0.467 to −0.003) and diabetes (β = −0.138; 95% CI: −0.258 to −0.017), whereas associations for hypertension and heart disease were not significant after socioeconomic adjustment. Among Hispanic participants, none of the total effects remained statistically significant in fully adjusted models (**Figure 4** and **Table S2**).

Sex-stratified analyses were generally consistent with the overall pattern (**Figure 4** and **Table S2**). In fully adjusted models, hypertension and heart disease were associated with lower cognitive performance in both males (β = −0.203 and −0.172, respectively) and females (β = −0.307 and −0.311, respectively; all p ≤ 0.008). For stroke, the association persisted among females in fully adjusted models (β = −0.456; 95% CI: −0.648 to −0.276; p < 0.001) but not among males (p = 0.320). For diabetes, the total effect remained significant among females (β = −0.206; 95% CI: −0.330 to −0.083; p < 0.001) but not among males (p = 0.450).

### Mediation of CVRF–cognition associations through accelerated biological aging

Causal mediation analyses showed that accelerated biological aging significantly mediated the associations between CVRFs and cognitive performance (**Figure 4** and **Table S2**). In the overall sample, all four CVRFs demonstrated significant indirect effects in fully adjusted models. The strongest mediation was observed for diabetes (ACME = −0.109; 95% CI: −0.146 to −0.076), with accelerated biological aging accounting for 88.5% of the total association between diabetes and cognitive performance. After accounting for the mediator, the direct effect of diabetes on cognition was no longer statistically significant (ADE p = 0.730), suggesting that the association between diabetes and cognitive performance was largely mediated by accelerated biological aging. Substantial mediation was also observed for heart disease (ACME = −0.065; 95% CI: −0.092 to −0.042), with 27.2% of the total effect mediated through biological aging, followed by hypertension (ACME = −0.044; 95% CI: −0.063 to −0.029; 17.1% mediated) and stroke (ACME = −0.044; 95% CI: −0.077 to −0.014; 13.7% mediated) (all ACME p ≤ 0.004). These findings indicate that accelerated biological aging represents an important pathway linking multiple cardiovascular risk factors to poorer cognitive performance.

Race/ethnicity–stratified analyses demonstrated substantial heterogeneity in mediation by accelerated biological aging (**Figure 4** and **Table S2**). Among Non-Hispanic White participants, accelerated biological aging significantly mediated the associations between all CVRFs and cognitive performance after socioeconomic adjustment, including hypertension (ACME = −0.047; 95% CI: −0.069 to −0.026), diabetes (ACME = −0.114; 95% CI: −0.157 to −0.074), heart disease (ACME = −0.065; 95% CI: −0.094 to −0.037), and stroke (ACME = −0.047; 95% CI: −0.087 to −0.010) (all p ≤ 0.012). Among Hispanic participants, accelerated biological aging significantly mediated the associations of hypertension (ACME = −0.029; p = 0.014), diabetes (ACME = −0.099; p < 0.001), and heart disease (ACME = −0.065; p = 0.004) with cognitive performance, whereas mediation of the stroke–cognition association was no longer statistically significant after socioeconomic adjustment (p = 0.068). In contrast, among Non-Hispanic Black participants, none of the mediation effects remained statistically significant in fully adjusted models (all p ≥ 0.068).

Sex-stratified analyses further revealed differential mediation patterns (**Figure 4** and **Table S2**). In fully adjusted models, accelerated biological aging significantly mediated the associations of hypertension and heart disease with cognitive performance in both males (ACME = −0.037 and −0.039, respectively; both p ≤ 0.002; proportion mediated = 18.3% and 22.7%) and females (ACME = −0.050 and −0.095, respectively; both p < 0.001; proportion mediated = 16.5% and 31.2%), with consistently larger mediated effects observed among females. For stroke, mediation through accelerated biological aging persisted only among females after socioeconomic adjustment (ACME = −0.055; p = 0.004; proportion mediated = 11.8%), whereas no significant mediation was observed among males (p = 0.134). For diabetes, accelerated biological aging significantly mediated the association with cognitive performance in both sexes (males: ACME = −0.097; females: ACME = −0.117; both p < 0.001). Overall, accelerated biological aging accounted for a substantial proportion of the associations between CVRFs and cognitive performance, with the magnitude and persistence of mediation varying across racial/ethnic groups and between males and females.

## Discussion

In this nationally representative sample of older U.S. adults from NHANES 2011–2014, we examined associations among major CVRFs, accelerated biological aging quantified using the PhenoAge algorithm, and cognitive performance. Three principal findings emerged. First, hypertension, diabetes, heart disease, and stroke were each associated with accelerated biological aging. Second, these risk factors were independently associated with lower cognitive performance. Third, accelerated biological aging mediated a substantial proportion of the associations between CVRFs and cognition, particularly for diabetes. The magnitude and persistence of associations and mediation effects varied across racial/ethnic and sex groups. These findings position multisystem biological aging as an intermediary linking cardiometabolic risk to late-life cognitive health.

Associations between CVRFs and accelerated biological aging were moderate in magnitude, with diabetes demonstrating the strongest relationship. Participants with diabetes exhibited substantially higher PhenoAge acceleration relative to those without diabetes, followed by heart disease, hypertension, and stroke. These patterns are consistent with established mechanisms through which chronic metabolic and vascular stressors contribute to systemic physiological decline^13,14^. Prior work in a nationally representative sample of U.S. adults aged 50 years and older demonstrated that a one-year younger PhenoAge was associated with approximately 7% lower mortality risk and 2% lower risk of incident chronic disease over two years of follow-up^22^. In this context, the degree of biological aging acceleration observed among individuals with cardiometabolic conditions represents clinically meaningful differences in late-life health trajectories.

CVRFs were also associated with poorer cognitive performance, with stroke demonstrating the largest association, followed by hypertension and heart disease. These findings align with extensive evidence linking vascular risk to cognitive impairment through mechanisms including cerebrovascular injury, microvascular dysfunction, and neuroinflammation, reinforcing the central role of vascular health in shaping cognitive outcomes in later life^30,31^.

Accelerated biological aging accounted for a substantial proportion of the CVRF–cognition associations, particularly for diabetes, for which approximately 89% of the total association with cognitive performance was mediated through biological aging. This large mediation proportion may partly reflect the inclusion of glycated hemoglobin (HbA1c), a key biomarker of glycemic regulation used in diabetes diagnosis, within the PhenoAge algorithm, introducing potential overlap between the exposure and the biological aging measure. Prior studies have also reported strong associations between accelerated phenotypic aging and diabetes risk and progression, suggesting that PhenoAge captures metabolic processes closely linked to diabetes^32,33^. Mediation proportions were more modest for hypertension, heart disease, and stroke, indicating that both systemic aging processes and disease-specific mechanisms contribute to cognitive outcomes. These findings demonstrate that clinically accessible measures of multisystem aging capture a meaningful pathway through which cardiometabolic risk relates to cognitive performance in older adults.

Stratified analyses revealed heterogeneity across racial/ethnic and sex groups. Associations between CVRFs, accelerated biological aging, and cognitive performance were more consistent among Non-Hispanic White participants and females, whereas several associations were attenuated among Non-Hispanic Black and Hispanic participants and among males after socioeconomic adjustment. These patterns parallel observations from studies of epigenetic aging measures demonstrating differential associations with cognitive trajectories across racial and ethnic groups^34^. Such heterogeneity may reflect differences in cumulative life-course exposures, structural inequities, and stress-related biological pathways that are not fully captured by current biomarker-based aging algorithms^10,35^. The observed variability in mediation across subgroups raises important questions about whether clinically derived aging measures equivalently represent aging processes in diverse populations^34^. These findings underscore the need for longitudinal and mechanistic studies to clarify subgroup variation in pathways linking vascular risk, biological aging, and cognitive decline.

These results have important public health implications. In an aging population with a high burden of cardiometabolic disease, identifying mechanisms that connect vascular risk to cognitive impairment is critical. Our findings suggest that accelerated biological aging constitutes one pathway through which cardiovascular and metabolic conditions influence cognitive health. Interventions targeting modifiable vascular risk factors—such as blood pressure control and glycemic regulation—may confer cognitive benefits not only by preventing clinical vascular events but also by attenuating systemic biological aging processes ^30^. Because biological aging reflects multisystem physiological dysregulation, integrating aging metrics into vascular and cognitive research may enhance risk stratification and provide scalable biomarkers for monitoring intervention effects aimed at reducing late-life cognitive decline^36^.

Several limitations warrant consideration. First, the cross-sectional design precludes establishing temporal ordering among cardiovascular risk factors, biological aging, and cognitive performance. Although the observed mediation patterns are consistent with a pathway from vascular risk to accelerated biological aging and subsequent cognitive impairment, longitudinal studies are needed to confirm temporal sequencing. Second, while hypertension and diabetes incorporated objective clinical measures, heart disease and stroke relied partly on self-reported diagnoses and may be subject to misclassification, which would likely attenuate observed associations^20^. Third, biological aging was quantified using the PhenoAge algorithm, which captures one dimension of multisystem physiological aging and is a well-validated predictor of morbidity and mortality. Complementary biomarkers—including epigenetic and proteomic measures—may capture additional pathways linking vascular risk to cognitive decline^10,37^. Finally, residual confounding related to life-course socioeconomic and structural determinants of health may contribute to observed subgroup differences^38^.

In conclusion, accelerated biological aging functioned as an intermediary linking cardiovascular and cerebrovascular risk factors to cognitive performance in a nationally representative sample of older U.S. adults. The substantial mediation observed—particularly for diabetes—highlights the role of multisystem physiological aging in vascular contributions to cognitive impairment. These findings demonstrate that cardiometabolic disease burden relates to cognitive health in part through systemic aging processes. Incorporating biological aging metrics into research on vascular cognitive health may improve risk stratification and inform strategies aimed at mitigating late-life cognitive decline.

## Author Contributions

J.J.L., T.Y., and A.J.L. designed the study. J.J.L., A.D., and T.Y. prepared the data, conducted analyses, and drafted the manuscript. J.J.L. generated the results and interpreted the findings. A.J.L. conceived, supervised, and critically revised the manuscript. All authors had access to the data and approved the final manuscript.

## Supplementary Material

Supplementary material is available with the online version of this article.

## Data Availability

The National Health and Nutrition Examination Survey (NHANES) data are publicly available through the Centers for Disease Control and Prevention at https://www.cdc.gov/nchs/nhanes/.

