## Supplementary Materials for "Vascular Risk Factors, Biological Aging, and Cognitive Performance in a National Sample of Older U.S. Adults"

**Table S1**. Associations of CVRFs with standardized PhenoAge acceleration and cognitive performance, overall and by race/ethnicity and sex, NHANES 2011–2014. Estimates are presented as β coefficients with 95% confidence intervals and p values. Model 1 adjusts for demographic factors. Model 2 additionally adjusts for socioeconomic factors.

**Table S2**. Mediation analyses of associations between CVRFs and cognitive performance via standardized PhenoAge acceleration, overall and stratified by race/ethnicity and sex, NHANES 2011–2014. Total effects, average direct effects (ADE), average causal mediation effects (ACME), and proportion mediated (%) are presented as estimates with 95% confidence intervals and p values. Model 1 adjusts for demographic factors. Model 2 additionally adjusts for socioeconomic factors.

**Figure S1**. Relationship between biological age (PhenoAge) and chronological age among participants in NHANES 2011–2014, overall and stratified by race/ethnicity and sex. Blue lines represent fitted linear regression lines. Pearson correlation coefficients (r) and corresponding p values are shown.

**Table S1**. Associations of CVRFs with standardized PhenoAge acceleration and cognitive performance, overall and by race/ethnicity and sex, NHANES 2011–2014. Estimates are presented as β coefficients with 95% confidence intervals and p values. Model 1 adjusts for demographic factors. Model 2 additionally adjusts for socioeconomic factors.

| | | Model 1: Demographic Covariates | | | | Model 2: Demographic + Socioeconomic Covariates | | | |
|---|---|---|---|---|---|---|---|---|---|
| CVRFs | Group | CVRFs and Standardized PhenoAge Acceleration | | Standardized PhenoAge Acceleration and Cognitive Score | | CVRFs and Standardized PhenoAge Acceleration | | Standardized PhenoAge Acceleration and Cognitive Score | |
| | | Estimate [95% CI] | p | Estimate [95% CI] | p | Estimate [95% CI] | p | Estimate [95% CI] | p |
| Hypertension | Overall | 0.390 [0.302, 0.478] | <2.2e-16 | -0.200 [-0.232, -0.167] | <2.2e-16 | 0.342 [0.255, 0.428] | 1.38e-14 | -0.129 [-0.160, -0.098] | 4.64e-16 |
| | Non-Hispanic White | 0.405 [0.287, 0.523] | 2.52e-11 | -0.203 [-0.247, -0.159] | <2.2e-16 | 0.348 [0.232, 0.464] | 5.42e-09 | -0.135 [-0.177, -0.092] | 6.08e-10 |
| | Non-Hispanic Black | 0.369 [0.140, 0.598] | 0.002 | -0.106 [-0.166, -0.046] | 0.001 | 0.344 [0.116, 0.572] | 0.003 | -0.051 [-0.104, 0.001] | 0.057 |
| | Hispanic | 0.277 [0.077, 0.477] | 0.007 | -0.163 [-0.235, -0.091] | 1.03e-05 | 0.269 [0.070, 0.469] | 0.008 | -0.108 [-0.168, -0.047] | 0.001 |
| | Male | 0.386 [0.258, 0.514] | 4.87e-09 | -0.183 [-0.227, -0.140] | 4.89e-16 | 0.334 [0.208, 0.460] | 2.50e-07 | -0.112 [-0.153, -0.071] | 1.01e-07 |
| | Female | 0.419 [0.292, 0.546] | 1.42e-10 | -0.205 [-0.251, -0.159] | <2.2e-16 | 0.371 [0.245, 0.496] | 8.76e-09 | -0.137 [-0.181, -0.093] | 1.49e-09 |
| Diabetes | Overall | 0.820 [0.731, 0.909] | <2.2e-16 | -0.211 [-0.246, -0.177] | <2.2e-16 | 0.759 [0.671, 0.847] | <2.2e-16 | -0.142 [-0.175, -0.110] | <2.2e-16 |
| | Non-Hispanic White | 0.835 [0.709, 0.961] | <2.2e-16 | -0.219 [-0.266, -0.173] | <2.2e-16 | 0.762 [0.638, 0.886] | <2.2e-16 | -0.150 [-0.195, -0.105] | 5.86e-11 |
| | Non-Hispanic Black | 0.797 [0.628, 0.967] | <2.2e-16 | -0.084 [-0.147, -0.021] | 0.009 | 0.775 [0.604, 0.946] | <2.2e-16 | -0.036 [-0.092, 0.019] | 0.202 |
| | Hispanic | 0.835 [0.663, 1.008] | <2.2e-16 | -0.162 [-0.240, -0.085] | 4.61e-05 | 0.834 [0.661, 1.007] | <2.2e-16 | -0.119 [-0.184, -0.054] | 0.000 |
| | Male | 0.794 [0.667, 0.920] | <2.2e-16 | -0.205 [-0.251, -0.159] | <2.2e-16 | 0.746 [0.622, 0.869] | <2.2e-16 | -0.131 [-0.174, -0.088] | 3.65e-09 |
| | Female | 0.894 [0.761, 1.026] | <2.2e-16 | -0.207 [-0.256, -0.158] | 2.83e-16 | 0.819 [0.688, 0.951] | <2.2e-16 | -0.144 [-0.190, -0.097] | 1.83e-09 |
| Heart Disease | Overall | 0.578 [0.483, 0.672] | <2.2e-16 | -0.195 [-0.229, -0.162] | <2.2e-16 | 0.497 [0.403, 0.591] | <2.2e-16 | -0.130 [-0.161, -0.098] | 9.89e-16 |
| | Non-Hispanic White | 0.572 [0.445, 0.700] | <2.2e-16 | -0.201 [-0.246, -0.156] | <2.2e-16 | 0.478 [0.352, 0.605] | 2.48e-13 | -0.136 [-0.179, -0.093] | 6.86e-10 |
| | Non-Hispanic Black | 0.652 [0.435, 0.870] | 6.97e-09 | -0.102 [-0.163, -0.042] | 0.001 | 0.614 [0.395, 0.832] | 5.44e-08 | -0.050 [-0.104, 0.003] | 0.067 |
| | Hispanic | 0.651 [0.413, 0.888] | 1.22e-07 | -0.147 [-0.220, -0.074] | 9.30e-05 | 0.622 [0.381, 0.863] | 6.00e-07 | -0.105 [-0.166, -0.043] | 0.001 |
| | Male | 0.422 [0.293, 0.552] | 2.54e-10 | -0.182 [-0.226, -0.138] | 9.05e-16 | 0.343 [0.214, 0.472] | 2.02e-07 | -0.114 [-0.155, -0.073] | 5.94e-08 |
| | Female | 0.807 [0.661, 0.953] | <2.2e-16 | -0.196 [-0.244, -0.149] | 1.43e-15 | 0.715 [0.569, 0.860] | <2.2e-16 | -0.134 [-0.179, -0.089] | 8.60e-09 |
| Stroke | Overall | 0.431 [0.284, 0.578] | 1.05e-08 | -0.209 [-0.241, -0.176] | <2.2e-16 | 0.317 [0.173, 0.462] | 1.81e-05 | -0.138 [-0.169, -0.107] | <2.2e-16 |
| | Non-Hispanic White | 0.466 [0.267, 0.665] | 4.87e-06 | -0.215 [-0.259, -0.171] | <2.2e-16 | 0.328 [0.132, 0.524] | 0.001 | -0.146 [-0.188, -0.104] | 1.86e-11 |
| | Non-Hispanic Black | 0.188 [-0.099, 0.474] | 0.200 | -0.106 [-0.165, -0.047] | 0.000 | 0.156 [-0.129, 0.441] | 0.285 | -0.051 [-0.103, 0.001] | 0.057 |
| | Hispanic | 0.362 [-0.091, 0.816] | 0.118 | -0.160 [-0.232, -0.089] | 1.27e-05 | 0.342 [-0.108, 0.792] | 0.137 | -0.106 [-0.166, -0.046] | 0.001 |
| | Male | 0.446 [0.218, 0.674] | 0.000 | -0.191 [-0.235, -0.148] | <2.2e-16 | 0.256 [0.029, 0.484] | 0.027 | -0.122 [-0.162, -0.081] | 6.42e-09 |
| | Female | 0.452 [0.249, 0.655] | 1.39e-05 | -0.215 [-0.261, -0.169] | <2.2e-16 | 0.381 [0.182, 0.580] | 0.000 | -0.143 [-0.187, -0.100] | 1.80e-10 |

**Table S2**. Mediation analyses of associations between CVRFs and cognitive performance via standardized PhenoAge acceleration, overall and stratified by race/ethnicity and sex, NHANES 2011–2014. Total effects, average direct effects (ADE), average causal mediation effects (ACME), and proportion mediated (%) are presented as estimates with 95% confidence intervals and p values. Model 1 adjusts for demographic factors. Model 2 additionally adjusts for socioeconomic factors.

| Model 1: Demographic Covariates | | | | | | | | | |
|---|---|---|---|---|---|---|---|---|---|
| CVRFs | Group | Total Effect | p | ADE | p | ACME | p | Proportion Mediated | p |
| Hypertension | Overall | -0.353 [-0.449, -0.259] | <2.2e-16 | -0.276 [-0.371, -0.184] | <2.2e-16 | -0.077 [-0.103, -0.055] | <2.2e-16 | 22.1 [14.8, 31.8] | <2.2e-16 |
| | Non-Hispanic White | -0.395 [-0.505, -0.289] | <2.2e-16 | -0.312 [-0.421, -0.212] | <2.2e-16 | -0.083 [-0.116, -0.054] | <2.2e-16 | 21.0 [13.5, 31.2] | <2.2e-16 |
| | Non-Hispanic Black | -0.148 [-0.319, 0.020] | 0.088 | -0.108 [-0.281, 0.054] | 0.202 | -0.040 [-0.080, -0.010] | 0.004 | 25.1 [-96.4, 143.5] | 0.092 |
| | Hispanic | -0.033 [-0.210, 0.135] | 0.696 | 0.013 [-0.160, 0.176] | 0.880 | -0.046 [-0.088, -0.015] | 0.004 | 36.0 [-622.4, 844.8] | 0.696 |
| | Male | -0.295 [-0.436, -0.157] | <2.2e-16 | -0.224 [-0.358, -0.091] | <2.2e-16 | -0.070 [-0.109, -0.036] | <2.2e-16 | 23.9 [12.1, 47.0] | <2.2e-16 |
| | Female | -0.402 [-0.527, -0.276] | <2.2e-16 | -0.317 [-0.437, -0.196] | <2.2e-16 | -0.085 [-0.125, -0.051] | <2.2e-16 | 21.2 [12.5, 33.4] | <2.2e-16 |
| Diabetes | Overall | -0.242 [-0.337, -0.143] | <2.2e-16 | -0.069 [-0.176, 0.032] | 0.178 | -0.173 [-0.218, -0.133] | <2.2e-16 | 71.2 [47.8, 123.0] | <2.2e-16 |
| | Non-Hispanic White | -0.256 [-0.383, -0.132] | <2.2e-16 | -0.071 [-0.190, 0.051] | 0.226 | -0.185 [-0.243, -0.131] | <2.2e-16 | 71.9 [45.4, 132.8] | <2.2e-16 |
| | Non-Hispanic Black | -0.238 [-0.393, -0.088] | 0.004 | -0.170 [-0.328, -0.007] | 0.040 | -0.068 [-0.133, -0.009] | 0.020 | 27.8 [3.0, 86.8] | 0.024 |
| | Hispanic | -0.138 [-0.324, 0.040] | 0.116 | -0.000 [-0.178, 0.180] | 0.970 | -0.137 [-0.218, -0.064] | <2.2e-16 | 90.3 [-488.3, 730.5] | 0.116 |
| | Male | -0.130 [-0.269, 0.009] | 0.078 | 0.032 [-0.106, 0.170] | 0.638 | -0.162 [-0.229, -0.103] | <2.2e-16 | 117.9 [-705.1, 772.3] | 0.078 |
| | Female | -0.350 [-0.498, -0.205] | <2.2e-16 | -0.166 [-0.313, -0.018] | 0.018 | -0.184 [-0.250, -0.124] | <2.2e-16 | 52.8 [33.7, 91.8] | <2.2e-16 |
| Heart Disease | Overall | -0.379 [-0.479, -0.278] | <2.2e-16 | -0.267 [-0.372, -0.166] | <2.2e-16 | -0.112 [-0.148, -0.083] | <2.2e-16 | 29.8 [20.3, 42.9] | <2.2e-16 |
| | Non-Hispanic White | -0.406 [-0.523, -0.287] | <2.2e-16 | -0.290 [-0.401, -0.179] | <2.2e-16 | -0.116 [-0.160, -0.077] | <2.2e-16 | 28.5 [18.9, 42.0] | <2.2e-16 |
| | Non-Hispanic Black | -0.167 [-0.349, 0.020] | 0.064 | -0.099 [-0.280, 0.088] | 0.268 | -0.068 [-0.128, -0.020] | 0.002 | 38.4 [-56.1, 254.3] | 0.066 |
| | Hispanic | -0.281 [-0.524, -0.033] | 0.032 | -0.184 [-0.427, 0.059] | d | -0.097 [-0.172, -0.036] | 0.002 | 33.8 [9.8, 142.0] | 0.034 |
| | Male | -0.302 [-0.436, -0.166] | <2.2e-16 | -0.226 [-0.354, -0.094] | <2.2e-16 | -0.076 [-0.120, -0.039] | <2.2e-16 | 25.0 [13.2, 46.8] | <2.2e-16 |
| | Female | -0.468 [-0.631, -0.309] | <2.2e-16 | -0.310 [-0.465, -0.148] | <2.2e-16 | -0.158 [-0.221, -0.100] | <2.2e-16 | 33.7 [21.3, 54.7] | <2.2e-16 |
| Stroke | Overall | -0.491 [-0.660, -0.328] | <2.2e-16 | -0.401 [-0.563, -0.241] | <2.2e-16 | -0.089 [-0.137, -0.045] | <2.2e-16 | 18.0 [9.6, 30.1] | <2.2e-16 |
| | Non-Hispanic White | -0.523 [-0.716, -0.334] | <2.2e-16 | -0.422 [-0.600, -0.241] | <2.2e-16 | -0.102 [-0.163, -0.047] | 0.004 | 19.2 [9.5, 32.9] | 0.004 |
| | Non-Hispanic Black | -0.329 [-0.597, -0.062] | 0.024 | -0.308 [-0.575, -0.043] | 0.032 | -0.021 [-0.061, 0.010] | 0.216 | 5.9 [-4.9, 30.9] | 0.220 |
| | Hispanic | -0.204 [-0.764, 0.358] | 0.474 | -0.144 [-0.703, 0.409] | 0.628 | -0.060 [-0.137, -0.001] | 0.046 | 14.2 [-270.8, 280.7] | 0.484 |
| | Male | -0.402 [-0.679, -0.127] | <2.2e-16 | -0.318 [-0.567, -0.050] | 0.004 | -0.085 [-0.168, -0.009] | 0.020 | 20.8 [3.3, 61.4] | 0.020 |
| | Female | -0.546 [-0.758, -0.331] | <2.2e-16 | -0.450 [-0.646, -0.242] | <2.2e-16 | -0.096 [-0.163, -0.036] | <2.2e-16 | 17.5 [7.3, 33.8] | <2.2e-16 |

| Model 2: Demographic + Socioeconomic Covariates | | | | | | | | | |
|---|---|---|---|---|---|---|---|---|---|
| CVRFs | Group | Total Effect | p | ADE | p | ACME | p | Proportion Mediated | p |
| Hypertension | Overall | -0.258 [-0.342, -0.177] | <2.2e-16 | -0.213 [-0.298, -0.131] | <2.2e-16 | -0.044 [-0.063, -0.029] | <2.2e-16 | 17.1 [10.2, 27.7] | <2.2e-16 |
| | Non-Hispanic White | -0.292 [-0.393, -0.198] | <2.2e-16 | -0.246 [-0.342, -0.148] | <2.2e-16 | -0.047 [-0.069, -0.026] | <2.2e-16 | 15.8 [8.4, 26.2] | <2.2e-16 |
| | Non-Hispanic Black | -0.070 [-0.208, 0.066] | 0.304 | -0.053 [-0.187, 0.083] | 0.454 | -0.017 [-0.042, 0.001] | 0.070 | 16.0 [-226.2, 289.8] | 0.350 |
| | Hispanic | -0.017 [-0.156, 0.114] | 0.838 | 0.012 [-0.125, 0.146] | 0.840 | -0.029 [-0.059, -0.006] | 0.014 | 20.2 [-666.3, 735.9] | 0.844 |
| | Male | -0.203 [-0.328, -0.085] | <2.2e-16 | -0.166 [-0.293, -0.052] | 0.006 | -0.037 [-0.062, -0.016] | <2.2e-16 | 18.3 [7.5, 43.8] | <2.2e-16 |
| | Female | -0.307 [-0.428, -0.190] | <2.2e-16 | -0.256 [-0.376, -0.145] | <2.2e-16 | -0.050 [-0.078, -0.026] | <2.2e-16 | 16.3 [8.5, 28.7] | <2.2e-16 |
| Diabetes | Overall | -0.124 [-0.208, -0.040] | <2.2e-16 | -0.015 [-0.104, 0.075] | 0.730 | -0.109 [-0.146, -0.076] | <2.2e-16 | 88.5 [45.9, 269.9] | <2.2e-16 |
| | Non-Hispanic White | -0.136 [-0.239, -0.038] | 0.018 | -0.022 [-0.129, 0.088] | 0.704 | -0.114 [-0.157, -0.074] | <2.2e-16 | 84.9 [40.6, 275.9] | 0.018 |
| | Non-Hispanic Black | -0.138 [-0.258, -0.017] | 0.030 | -0.110 [-0.251, 0.030] | 0.118 | -0.028 [-0.078, 0.019] | 0.270 | 19.1 [-20.3, 145.8] | 0.296 |
| | Hispanic | -0.033 [-0.173, 0.101] | 0.652 | 0.066 [-0.069, 0.211] | 0.398 | -0.099 [-0.164, -0.039] | <2.2e-16 | 94.9 [-1622.9, 2005.8] | 0.652 |
| | Male | -0.045 [-0.160, 0.075] | 0.450 | 0.051 [-0.074, 0.171] | 0.432 | -0.097 [-0.148, -0.051] | <2.2e-16 | 113.2 [-2438.9, 1432.8] | 0.450 |
| | Female | -0.206 [-0.330, -0.083] | <2.2e-16 | -0.089 [-0.215, 0.032] | 0.172 | -0.117 [-0.173, -0.067] | <2.2e-16 | 57.7 [29.9, 131.9] | <2.2e-16 |
| Heart Disease | Overall | -0.237 [-0.327, -0.149] | <2.2e-16 | -0.173 [-0.268, -0.081] | <2.2e-16 | -0.065 [-0.092, -0.042] | <2.2e-16 | 27.2 [15.7, 47.8] | <2.2e-16 |
| | Non-Hispanic White | -0.265 [-0.369, -0.169] | <2.2e-16 | -0.201 [-0.303, -0.094] | <2.2e-16 | -0.065 [-0.094, -0.037] | <2.2e-16 | 24.1 [13.3, 43.2] | <2.2e-16 |
| | Non-Hispanic Black | -0.068 [-0.249, 0.115] | 0.472 | -0.038 [-0.214, 0.147] | 0.712 | -0.030 [-0.069, 0.001] | 0.068 | 21.2 [-455.2, 522.1] | 0.492 |
| | Hispanic | -0.093 [-0.276, 0.095] | 0.346 | -0.028 [-0.214, 0.169] | 0.806 | -0.065 [-0.119, -0.018] | 0.004 | 43.0 [-712.4, 732.3] | 0.350 |
| | Male | -0.172 [-0.301, -0.045] | 0.008 | -0.133 [-0.262, -0.013] | 0.036 | -0.039 [-0.068, -0.016] | 0.002 | 22.7 [7.6, 74.2] | 0.010 |
| | Female | -0.311 [-0.446, -0.173] | <2.2e-16 | -0.216 [-0.351, -0.085] | <2.2e-16 | -0.095 [-0.146, -0.051] | <2.2e-16 | 30.7 [16.0, 55.3] | <2.2e-16 |
| Stroke | Overall | -0.317 [-0.471, -0.165] | <2.2e-16 | -0.273 [-0.435, -0.119] | <2.2e-16 | -0.044 [-0.077, -0.014] | 0.004 | 13.7 [4.7, 30.2] | 0.004 |
| | Non-Hispanic White | -0.334 [-0.514, -0.168] | <2.2e-16 | -0.287 [-0.462, -0.109] | 0.002 | -0.047 [-0.087, -0.010] | 0.012 | 13.9 [3.0, 33.8] | 0.012 |
| | Non-Hispanic Black | -0.238 [-0.467, -0.003] | 0.048 | -0.230 [-0.455, 0.007] | 0.058 | -0.008 [-0.029, 0.008] | 0.350 | 2.4 [-6.1, 20.6] | 0.374 |
| | Hispanic | -0.142 [-0.556, 0.274] | 0.514 | -0.107 [-0.515, 0.317] | 0.630 | -0.036 [-0.085, 0.002] | 0.068 | 10.8 [-191.7, 243.4] | 0.562 |
| | Male | -0.131 [-0.382, 0.109] | 0.320 | -0.100 [-0.348, 0.145] | 0.422 | -0.031 [-0.081, 0.009] | 0.134 | 15.9 [-265.2, 220.8] | 0.382 |
| | Female | -0.456 [-0.648, -0.276] | <2.2e-16 | -0.402 [-0.590, -0.219] | <2.2e-16 | -0.055 [-0.102, -0.019] | 0.004 | 11.8 [3.9, 24.9] | 0.004 |

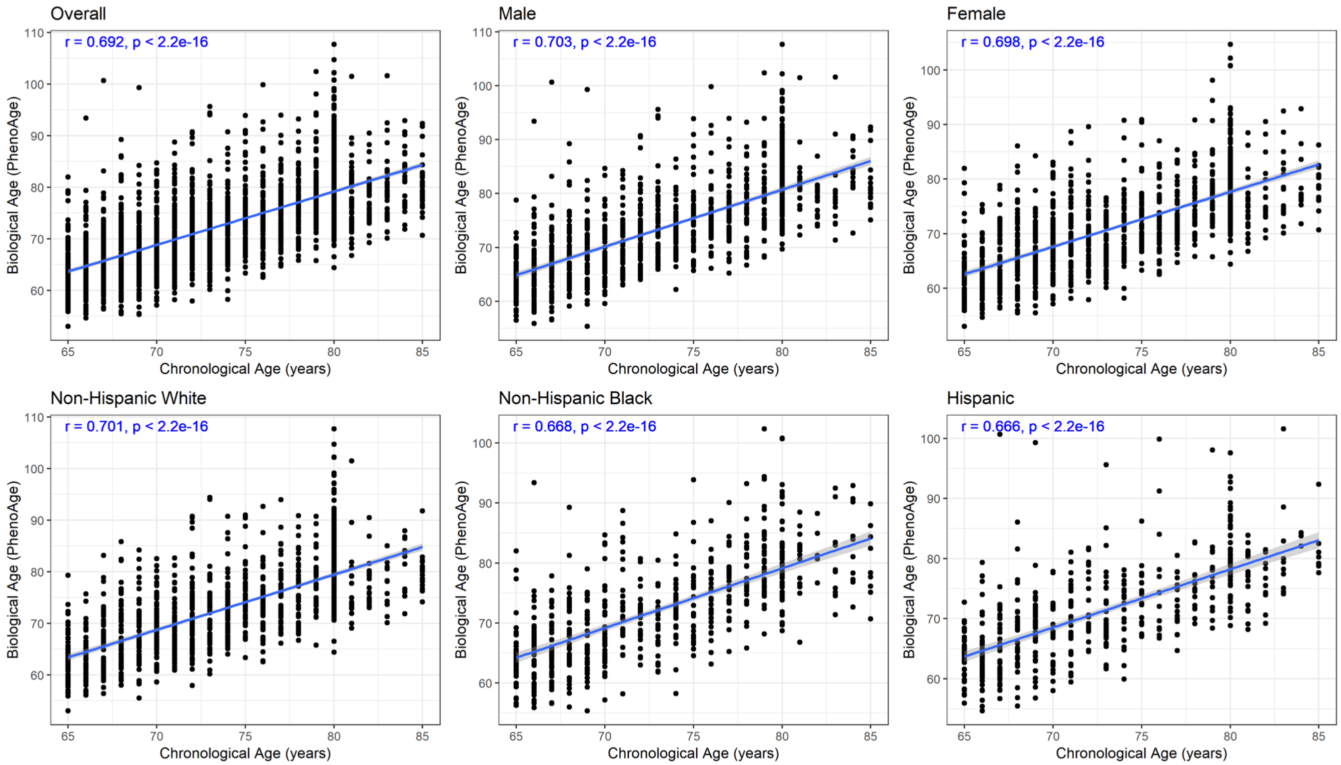


**Figure S1**. Relationship between biological age (PhenoAge) and chronological age among participants in NHANES 2011–2014, overall and stratified by race/ethnicity and sex. Blue lines represent fitted linear regression lines. Pearson correlation coefficients (r) and corresponding p values are shown.
